# Investigating How East Asian Medical and Biomedical Systems Can Communicate: A Qualitative Exploration of How East Asian Medicine Concepts Can be Used in Modern Health Research

**DOI:** 10.1101/2024.01.14.24300930

**Authors:** JoHannah Macbeth, Lisa Conboy, Maria Engels, Rachel Burack

## Abstract

**Purpose:** There is much published science supporting East Asian Medicine (EAM) as a treatment for many health issues. Public Health has made great advancement in understanding the effects of health disparities globally. East Asian Medical Models offer additional ways to think about health disparities.

**Methods:** Twelve participants who are Biomedicine and/or Eastern Medicine experts or social scientists were identified from the research team’s network. Our goal in this purposive sample was to represent diverse backgrounds, educational and professional experience, expertise, age, gender, and geographic location. Following IRB approval, all potential subjects identified for recruitment agreed to participate and were consented. Semi-structured interviews were conducted with each subject on the Zoom © platform, and ranged between 60 and 90 minutes. Recordings were transcribed and checked for accuracy, and double coded for themes. Any disagreement in coding was addressed in discussion between the coders. We assessed the data for saturation on key content areas, and when finding repetition of themes, chose to complete the stage of data analysis. Next, we employed a thematic analysis to better understand the key concepts.

**Results:** Seven main themes emerged from the interviews. These are necessary areas to consider when considering health disparities: THREE TREASURES, PATIENT PROVIDER RELATIONSHIP, ASSESSMENT OF HEALTH STATUS in EAM, PRENATAL ESSENCE, POSTNATAL ESSENCE, DETERMINANTS OF HEALTH, BELONGING.

**Conclusions:** Scientific discovery and health care will improve with improvement in communication between medical practitioners and scientists from EAM and Biomedicine (BM). We use our data to consider ways that the EAM model of health can add to our understanding of the effects social disparities on health.

## INTRODUCTION

The contemporary scientific study and clinical use of acupuncture are growing. A recent search of Pubmed for the terms ‘East Asian Medicine”, “Chinese Medicine,” or “Acupuncture” offered over 350,000 citations. With notable exceptions little clinical science has been published focusing on the translation of core medical terms across systems of medicine. Some ideas innovative to Biomedicine (BM) have been established within East Asian Medicine (EAM) for thousands of years (e.g. interstitium which is the Sanjiao channel). Improving communication between medicines will accelerate more discovery. Through deeper understanding of what already exists, new advances and true discovery can occur and ultimately improve the health of the public.

This research uses stakeholder voices to encourage a greater understanding between EAM and BM systems of medicine. Using data from focused interviews with 12 stakeholders we explore the EAM theory of health and the factors influencing health. Together with our interviewees we examine how EAM theory conceptualizes the interplay of inherited factors, with lived conditions and experiences.

## BACKGROUND

EAM and BM have both shared and divergent ideas of what health is, and how to improve the lived experience of patients. The World Health Organization (WHO) defines the determinants of health to include (DOH): *“the social and economic environment, the physical environment, and the person’s individual characteristics and behaviors (CDC, 2023)”*. Much modern epidemiologic science attempts to explain how these contextual conditions that exist outside the individual patient can affect the body. While this work has led to reduced suffering, there is still a lack of theoretical understanding of some of the complexity of the actions of DOH, such as why illness affects different types of people differently, and how context, such as the therapeutic interaction, can ameliorate the negative effects of such identified DOH.

EAM is a whole system of medicine which uses a model of health and disease that offers a *“non pharmaceutical treatment, simple application, wide range of use, good curative effect, and low cost” (*Xinnong*, pg. “foreword”).* The EAM model of health and disease includes many of the same DOH found in epidemiologic science.

> On the basis of previous literature, it takes the theories of yin-yang, five elements, zang-fu, meridians and collaterals, mentality and spirit, qi and blood, body fluid, five emotions and six exogeneous (sic) pathogenic factors as the basic knowledge of traditional Chinese medicine, and acupuncture and moxibustion as the main therapeutic technique; it explained the physiology and pathology of the human body, the principles of diagnosis, the prevention and treatment of diseases from the perspective of atheism, holistic conception, the viewpoint of development and change, and the relationship between the human body and the natural environment. This laid a theoretical foundation of Chinese medicine and pharmacology, including acupuncture and moxibustion. (CAM, pg. 3).

Based on EAM theory, there are many routes of therapeutic intervention. A wide set of potential influencers on health and illness, together with a commensurate wider set of ways to intervene for patients’ health, support novel scientific questions of how we can reduce the effects of negative DOH.

EAM situates the human ‘between heaven and earth’, modeling the human at the center in relation to the patient’s spirit, the relation to the patient’s own biology, and the relation to the patient’s environment (Beinfield, H., & Korngold, E. 1992). EAM theory is built on core constructs, the *Three Treasures* termed *Jing* (Essence)*, Qi,* and *Shen* (Mind) (see *Figure 1)*. In EAM, “Essence, *Qi* and Mind also represent three different states of condensation of *Qi*, the Essence being the densest, *Qi* being more rarefied, and the Mind being the most subtle and immaterial. According to Chinese medicine, Essence and *Qi* are the essential foundation of the Mind.” (Macocia, pg. 50). *“If Jing and Qi are strong, the Shen will be happy*”. (Subject 1) The *Three Treasures* are the foundation from which clinical practice is built.

The authors *(R.B., M.E., J.M., L.C. - see Appendix 1*) are acupuncture providers or scientists who proposed this research project to consider the question of whether generational low income/ socioeconomic status is related to or can be understood to result in less inherited *Jing*? *Jing* (or *Essence*) is both inherited and accumulated. Inherited *Jing* occurs at fertilization of the egg by the sperm (similar to inherited genetics). Accumulated *Jing* occurs throughout life and is intimately linked with socioeconomic factors (epigenetic expression). We discovered, however, in speaking with our experts that the story was more complex than a single answer to this question, and that we needed to consider *Jing* in the context of the holism of EAM theory.

This idea for this research began during the first year of the COVID-19 pandemic, with a worldwide increased interest in health, the factors that influence health, and socially-caused incongruities in health. We wanted to learn from our stakeholders whether EAM theory can add to our understanding of BM, epidemiology, and enrich our healthcare system. Our interviewees reminded us that the DOH are always considered in the context of the patient’s experience and are clinically found through the analysis of the *Three Treasures*.

## METHODS

The Institutional Review Board at Massachusetts College of Pharmacy and Health Sciences oversaw the data collection process (IRB011521C). The Institutional Review Board at Seattle Institute of East Asian Medicine oversaw the data analysis process (#9-17-22).

Twelve participants who are BM and/or EAM experts or social scientists were identified from the research team’s network. Our goal in this purposive sample was to represent diverse backgrounds, educational and professional experience, expertise, age, gender, and geographic location. All potential subjects identified for recruitment agreed to participate. De-identified information about each of the final interviewees is in *Appendix 2* (see attached.)

Interviews were conducted and recorded virtually using the Zoom © video conferencing platform. The consenting process began with a brief introduction to the team, and a conversation about the purpose of the research project, and instructions/assurances regarding confidentiality. Consent was requested and given by each participant at the beginning of each interview. Each interviewer, either author R.B., M.E., or J.M., took turns leading interviews by using a scripted introduction and image, asking seven interview questions and using probes to follow-up (*see Appendix 3 for consent document and Appendix 4 for research questions)*. Throughout the interview the remaining two interviewers took notes on areas they found of interest or relevance.

Recordings were transcribed and checked for accuracy. Transcripts were read at least twice by two authors (either R.B., M.E., or J.M.) and then double coded for themes independently. Coding consisted of extracting sought and emergent themes as well as condensing responses to each queried topic across the sample. Any disagreement in coding was addressed in discussion between the coders. Data was assessed for saturation on key content areas, and when finding repetition of themes, chose to complete the stage of data analysis. Next, we employed a thematic analysis to better understand the key concepts.

Due to the nature of the semi-structured interview agenda, measuring frequencies of the qualitative reporting of particular themes made little sense. Instead, terms such as ‘all’, ‘most’, ‘a few’ and ‘one’ were used following the recommendations of Price et al.

## RESULTS

All 12 members of the target sample agreed to participate. Resulting interviews ranged from 60 to 90 minutes. Our seven resulting themes and definitions are explained below using the exemplary quotes from our research. These exemplary quotes illustrate the common values and expertise conveyed by the research respondents and the resulting data. More examples of quotes for each theme can be found in Table 2.

### THREE TREASURES

Definition: *A representation of three factors referenced in East Asian Medicine as Jing, Qi, and Shen. The unique expression of an individual’s core nature which allows the determination of health to be made, influenced by prenatal (genetics) and postnatal (epi-genetics) factors*.

*Jing* is the inherited Essence from both parents. It is similar to the biomedical idea of genetics yet differs because it is recognized as also the inherited Essence of all the previous generations captured at the moment of conception. *Qi* and *Shen* are comprised of inherited *Jing* (genetics) but act similarly to epigenetics with influences post-birth including but not limited to access to food, safety, belonging, etc. being important to how it presents. When asked about the relationship of the *Three Treasures* respective to genetics and epigenetics, one participant reconfirmed these relationships with additional discussion of genetic heritage and the postpartum time frame:

> So it’s, yes, it’s Qi and blood and Jing. … the Three Treasures, … I think to disregard the potential, the influence of history and the potential reality of future is, in my mind, a little short sighted. (Subject 6: 12:48)

The *Three Treasures* are the basis of existence. In EAM, living in harmony with one’s nature (e.g. eating with the seasons) is protective and sustainable.

> Jing is qi, and qi is qi. So if you’re living a balanced life, by definition, you are not depleting it. (Subject 1:1:04:05)

Other respondents tell us about the interplay and interdependence of the *Three Treasures* and how their interactions support the individual’s engagement in the world.

> Jing nourishes or fortifies the body and the Qi will mobilize it. But, the Shen engages the body in the world. (Subject 12: 41:10)

Our respondents told us about the *Three Treasures,* and how this concept is used differently from similar biomedical ideas. Having moderation with lifestyle behaviors supports an individual of their *qi* and *jing*. The composition of *Three Treasures* informs an individual’s state of mind, toward happiness, resilience, ability to evolve, etc.

### PATIENT PROVIDER RELATIONSHIP

Definition: *The relationship between patient and provider must be professional and intentionally therapeutic. Patients are seen as whole persons and health-related goals are created in a collaborative and mutual manner to optimize outcomes*.

Many participants referenced the importance of and cultivation in self knowledge and its relationship to *Jing* and socioeconomic disadvantage. This is true for both patient and provider as they work together for the patient’s health. Not only is the individual a reflection of their biosphere but they are also a personification of their experiences; this understanding is integral to the application of the medicine. A provider’s ability to be self-reflective (or the lack thereof) will support (or inhibit) their ability to listen to a patient.

> A large portion of the actual treatment and the success of a treatment is listening to a person’s story. You don’t need to comment on the story. But you need to listen to it. You need to keep boundaries around the story, you need to keep them tight, whatever your comfort level is… you need to listen to them from your first encounter… People want to tell you their stories, and you need to listen. We can be really powerful mirrors of people’s lives, to help them navigate and make change. (Subject 1: 26:17).

EAM is a whole system of medicine, and healthcare, as well as a way of life. Self-cultivation by both the patient and practitioner are encouraged and embedded within the mentality of life-long learning. In the EAM model, the patient provider relationship is influenced by the *Three Treasures* of the patient, the provider, and their consequent relationship. Clinical outcomes are determined by both the patient’s *and* the provider’s *Three Treasures*.

In the EAM model, the established patient provider relationship encourages autonomy of the patient that is quite unique and novel to medicine. Instead of the provider holding all of the answers, the provider is the conduit for the patient’s healing - ultimately learning from the best possible teacher: the patient.

> Respecting one’s boundaries [is the most important factor in building a relationship]. (Subject 9: 41:35)

Each clinic session is a unique opportunity for healing, created by a dynamic patient, dynamic provider, in a dynamic context, all of which can change interaction to interaction. Our research emphasized the importance of self-cultivation especially as a healthcare provider in order to reach shared healthcare goals established by the patient and provider. The EAM collaborative patient provider relationship model puts the patient above the provider and views the patient as the expert of their own health. Many of our respondents told us that having one trusted support system, in this case - the healthcare provider, could help ameliorate aspects of social-economic disadvantage, thus adding a protective factor to the *Three Treasures.* (Subjects 5 and 7).

### ASSESSMENT OF HEALTH STATUS IN EAM

Definition: *The collaborative process between the patient and provider where the provider gathers information using all five senses, integrates this information in the foundational framework of the body systems (10 Key Questions /Appendix 5) viewing the patient holistically; mentally, emotionally, physically, and spiritually*.

The Assessment of Health Status code relies on the theory of EAM, including the integration of information gathered by the practitioner using the five senses to accurately assess the health status of the patient.

> Body language [**of the clinician**] is huge [**significant factor**] to actually be open versus closed. If we are open, and have an open heart with the patients, I think they will feel that they have our attention, respect, and understanding to treat them. (Subject 5: 1:05:45)

Respondents highlighted the importance of listening, observation, BM, eastern science, inner science, and individualized health care (see Table 2) when discussing an Assessment of Health Status. The 10 Key Questions framework for EAM allows for the patient’s lived experiences to have influence in the eventual diagnosis - as in, a thorough examination of the patient is administered, looking for potential links that would not stand out within another medicine’s framework. The 10 Key Questions assess the state of the *Three Treasures* in order to come to a diagnosis.

> Biomedicine is very colonial – ‘scientific’ discounts a lot of important science and negates the relevance of other systems, which means we are missing out on some important opportunities that [ultimately] impact medicine and public health. (Subject 3: 7:00).

> Predisposition is sometimes discussed within Biomedicine in a manner that actually dismisses the impact of humans. Like racism, black mothers and death/mortality rates, etc. But does not focus on the impact from other humans: impact of being enslaved and that stress impact for generations, etc. Seems like a lot of emphasis on the focus of ‘here and now’ of predisposition without really discussing/honoring the past wounds that led to the now. (Subject 3: 17:24).

The Assessment of Health Status can capture these various dynamics that can impact the ultimate starting point for optimal health: patients want to be heard, to be understood, and to feel a sense of belonging.

### PRENATAL ESSENCE

Definition: *Fixed at birth, this is the accumulation of contextual and physical influences from parents, fetal experiences while in utero, and generational factors. (See Figure 1 for more detail.)*

The physical, social, emotional, and spiritual environments significantly influence the Prenatal Essence of each individual. Knowing one’s generational history is important to knowing who you are today. Generational patterns develop within cultural traditions and are cumulative in effect. Examples include access to certain natural resources, education, political positions, or lifestyle. Conversely, lack of access or societal impacts like racism, poverty, war, or famine can lead to the development of generational trauma patterns. Interviewees shared these ideas above and concluded that the concept of Prenatal Essence in EAM can partially find its synonym within the understanding of genetics.

As one interviewee suggested, the understanding of prenatal Essence “*captures the idea of energetic potential; this concept does not necessarily get captured that way within [**bio**]medicine.“* (Subject 7: 12:25).

> Pre [**Natal Essence**] is…the coming of our ancestors together to create. So it’s not just the mother and father, but it’s the whole ancestry all coming together. (Subject 6: 6:09)

> [**PreNatal Essence**] is responsible for growth and maturation and reproduction and a whole bunch of things. Right? It’s the strength of constitution. It’s our Western genetic heritage, if you will. (Subject 6: 5:15)

An individual living in the present day is truly the culmination of their history and their family history. Cultural exposures, even generations back, influence the function, quality, and amount of prenatal Essence. With modern scientific advancement of BM in genetics and aiding the conception process (i.e In vitro fertilization-IVF), interesting questions now arise such as can this generational inheritance of *Prenatal Essence* be influenced, and on what scale? One interviewee explains how EAM ideas can integrate with contemporary biological understandings if we consider the importance of context:

> It’s what we do know, that the carrier mom, the “matriarchal carrier”, I think is the term, influences the micro RNA of the fetus, which changes their post[**natal Essence**]. (Subject 6, 45:19)

Other practitioners share a similar point:

> In the older texts, they would say that the prenatal [**Essence]** you really can’t [**change**], it’s generational… But now, maybe we can change it with genetic manipulation. (Subject 12, 6:29)

For healthcare providers, a logical starting point to consider is the understanding of *jing* (prenatal) deficiency. What if the fetus doesn’t have enough *jing* and is therefore unable to fight from this disadvantaged state? At the very basic cellular level, life is designed to live. Within the structure and design of prenatal Essence we were reminded of the inseparable context of prenatal Essence within the *Three Treasures*.

> Jing… is never isolated from the three treasures. (Subject 12: 51:49)

This interconnection of forces was also examined to consider the effects of trauma. One interviewee talked about the influence of trauma:

> However, I will say that based on what I’ve learned about trauma, and the impacts of trauma, (slight pause) trauma is real, and the impact of trauma is real. And, it can influence generations. So the trauma that one experiences could impact one’s offspring, even if that offspring hasn’t experienced the same trauma. So we talk a lot about generational trauma. And there’s a lot of [discussion that in reality] holistic care for children doesn’t start when they’re born, it starts when the mother is pregnant, because that whole experience in the womb has an impact when the child is born. (Subject 4, 12:01)

Other practitioners shared a similar perspective:

> Humans are made to adapt. So, there’s always adaptations to our environment, right? What we don’t recognize is whether that adaptation happens within the natural environment or within manmade structures… Where people are either allowed to live free or live within oppressive societies or structures… The changes or the adaptations that happen, really [become pronounced]… This is a challenging subject, because this is a perfect example where [bio]medicine just continues to ignore chronic issues. One of the most telling issues is when we look at the impact of stress on mammals… They did a study on the infant mortality rates, and pregnancy mortality rates of women, black women in particular. (Subject 3:12:46)

Because prenatal Essence *(Jing)* works synergistically and dependently, it will borrow from the other two treasures when necessary in pursuit of physical, social, and energetic homeostasis. Balance between the *Three Treasures* is necessary to their working and sustained dynamics as a system.

### POSTNATAL ESSENCE

Definition: *The accumulation of contextual and physical influences from parents/caregivers, patient experiences, and generational factors post-birth*.

Postnatal Essence is more influential and a foundational construct in EAM. Postnatal Essence focuses on how each individual can integrate lifestyle practices to nourish and cultivate their own vital life force. As one practitioner stated:

> We can’t necessarily change the genetics, but the epigenetics can certainly be influenced. (Subject 12: 16:11)

Just as prenatal *Jing* exists in a context of the *Three Treasures*, so postnatal *Jing* exists in a context of the *Three Treasures*. There is just more opportunity for the individual to influence the balance of their specific *Three Treasures* on a day to day basis, with lifestyle choices.

> Pre[natal] Essence (Jing) is very hard to deplete; [however,] post[natal Essence] (Jing) [can be more easily depleted]. [Clinically it is] very hard to distinguish [between] the two, so I don’t. (Subject 1: 11:47)

One interviewee discussed the influence of birth context, genetic and cultural background of both mother and baby.

> At birth, for a vaginal birth, the child is colonized with the mother’s vaginal flora which colonizes their gut for digestion. So if we think about it, in the terms that we’re talking now, that’s the mother’s, mother’s, mother’s, mother’s, mother’s, mother’s, mother’s, vaginal flora. I have this (post C-section) case right now (in Europe) with this child [who] has severe eczema. If the vaginal flora is healthy, if there’s no strep B, or HPV or any other disorder, they actually do a swab inside the cheek, the neck, under the arms, and the groin, the inguinal ligament area, and they found that there are less food allergies when they do that. Now we don’t do that in America and I don’t know why… So this child, what’s interesting is the mother is Filipino, and the father is Colombian. So it’s this beautiful kind of cultural mix. So we had the baby, the mom is cooking the baby traditional Filipino food, and we did a vaginal swab. Right, so the eczema started clearing because that Flora changed, right? And so that kind of process of honoring that kind of millennial, a millennia millennia of birth, and carrying on, has allowed this five month old baby to reset. Yeah. You’re talking about the influence of post[natal Essence]. (Subject 6, 18:15)

The term epigenetics can be likened to the Postnatal Essence that is identified in EAM. Individuals have direct impact and influence on their Postnatal Essence during their lifetime. One interviewee compared Postnatal Essence to having a bank account of health: one can invest and also deplete their account.

When asked how to evaluate and “see” *Jing,* one practitioner noted that it is important to be able to detect how much postnatal *Jing* an individual has in order to pursue a healthy life:

> Look at the face, where all the water places are that relate to Jing. Under the eyes, and by the chin. If you have a very strong energetic yang, you’ll see lines across the forehead, etc. Look at how fire yang someone is, how much earth is there. It’s not easy. This correlates Jing to water - AND Jing is primarily associated with water and looks at it through the water element. Not just water. It is where the kidneys come into play: they hold it, put it into place, etc. like the placenta and amniotic fluid, etc. (Subject 2: 19:08)

The potential influences on postnatal Essence are endless. The realms of socioeconomic status, access/lack of access, specific ideologies, environmental climates, educational opportunities, beliefs, or other health cultivating opportunities are recognized for being either the building blocks or destroying measures for the cultivation of the postnatal Essence. Providers and patients alike are encouraged to cultivate a lifestyle that supports the regeneration, protection, and nurturance of their postnatal Essence.

> Epigenetics is the most important factor that [affects] your health. (Subject 11: 6:05)

In EAM the influence of previous generations creates an “epigenetic blueprint” in all of us. The Essence of the human can help them rise above the proscriptions and norms of society. Our creative ability to cultivate the *Three Treasures* determines our mindset, behaviors, and outlook. Our ability to create is affected by context, including socioeconomic status. This creation will then ripple into the Essence passed to the next generation. (See Figure 1).

> Socioeconomic status informs all kinds of things. It informs the kind of opportunities you might have or not have in this world, it informs your sense of self. It ties in with family because you know, families have their story of who [they] are… Socioeconomic status is [determines] if you’re able to get educated to the level that’s commensurate with your intelligence, it’s going to have a lot to do with the food that you eat, [and] the people that you’re around. All of [these factors are] going to have a tremendous impact on what, I’m going to call, epigenetics. Epigenetics, as I think about it, [represent] the influences of this world that interact with the Essence of who you are and [either] give you an opportunity to flower parts of you or maybe even cut off parts of you. So yes, I would say that [in the EAM tradition]…all interact, interpenetrate and, influence each other simultaneously. (Subject 8: 27:10)

And similarly,

> I suspect that our family of origin is like our original epigenetic blueprint, in some ways, it gives us filters, it gives us…belief systems; what’s possible [and] what’s not possible. So, I think our original epigenetic kind of blueprint, in some ways, comes from our family of origin. (Subject 8: 36:37)

Prenatal and postnatal Essence are interdependent with *Jing, Qi,* and *Shen*. They are the *Three Treasures*, which are innately generative and sustainable. Evidence of the state of the postnatal Essence within the context of the *Three Treasure*s can be seen in various locations of the body. Diagnostic tools such as Face Reading use this method.

> When I’m reading the face, and I see someone’s fertility (Three Treasures), I’m looking at a really small area. I’m looking to see how wide the philtrum is, how long the philtrum is, how deep the philtrum is, and looking to see whether or not there’s the ability for the body to get pregnant, to hold that pregnancy, to maintain that pregnancy. It’s something that’s really, really important. (Subject 2: 12:28)

Postnatal Essence is the culmination of parents/caregivers, patient experiences, and generational factors post-birth. Prenatal and postnatal *Jing* exist in the context of the *Three Treasures* and that interplay is constant and dynamic.

### DETERMINANTS OF HEALTH (DOH)

Definition: *The factors that interplay and create a multi-dimensional impact on the mental, emotional, physical and spiritual health of an individual*.

Human beings are influenced by many components which can help or hinder moving towards optimal health. Components can include religion, socioeconomic status, geographic environment, zip code, ethnic identity, a history of trauma, access to education, or the amount of stress an individual is facing. With changes in context, a positive factor can become harmful.

> Same factors listed as positive or negative – just switched due to access. (Subject 4: 30:55)

Our patients live in an environment that affects which DOH they are exposed to and how those determinants affect them.

> I think that if you were born in a difficult situation with regards to the location that you’re born into, and with your abilities from that location, if you’re born in a very tough inner city neighborhood, for instance, then that is so hard to break out of from a generation standpoint, because of education; there’s such a disparity, especially now in the [COVID] pandemic, from one area to another. My wife works at an inner city school in Providence, RI, and there was a student who couldn’t get online because she couldn’t get her computer to connect, and she didn’t know what to do. So during the pandemic, my wife actually had to go visit her house… and they open up the laptop, and the girl says, “See, I don’t know, I type in the address and it doesn’t work.” My wife realized [the family] doesn’t have internet [in the home]. [The school] had to get a hotspot and set [the student up to be able to access the internet from home.] So, what [an educational] disadvantage here for someone that has no internet [to learn from home], and they’re living in a situation [where] mom and dad are away, mom and dad have to go work just to make ends meet… And this impacts nutrition, too. (Subject 5: 51:43)

On the other end of the spectrum, there can be too much parental support:

> One of the things I’ve seen with the college students [I teach] is that mom and dad have done so much, they’ve done too much. So now you don’t know how to operate on your own. And you don’t know how to navigate the world with other people who are from different cultures, because you have been isolated in a bubble, and that can also add to issues… So from the highest level, generational wealth is getting passed down, for the most part. I mean, some are choosing not to do that, but many are, and if you’re not passing the wealth down, for instance, then that child will [likely] still have [access to a] higher level of education and opportunities to be able to get into the schools and do those things so that they can be better off. (Subject 5: 54:35)

Social and economic patterns create situations with inertia for cycles that are difficult to break.

> So there is this perpetual wheel of motion that’s in play and very hard [to change] that’s going from generation to generation… also, [the child will mirror the behaviors and lifestyle of the parents, which further adds barriers to breaking free], and it does impact the overall health status from one generation to another. (Subject 5: 51:43)

Regardless of socioeconomic status humans all experience the struggle to survive.

> Stress of money and time impact both the upper echelons of society and low income. (Subject 1: 20:54)

Understanding the lifestyle context of when, where, and how a person was born is critical for clinical assessment, care, and the determination of optimal treatment outcomes. It is also important that the patients’ own considerations and assessments are integrated in the treatment plan.

> Patient’s self-reported health is on the list of how to identify the state of health. Some things aren’t going to show up on an MRI (or access to MRI) but they are feeling something. Not the best use of science, knowing if you have a gene because it doesn’t actually determine the health outcome, so many other things determine health outcomes (again, genetic roulette of what is expressed). (Subject 7: 24:36)

It often takes a lot of convincing, research, reminding, and habit forming skills to help an adult make positive lifestyle changes that will develop the best possible epigenetics. In contrast, children are living in the context of their own rapidly developing minds and bodies. For an adult:

> Genetics come out faster, and epigenetics comes out slower. (Subject 2: 31:12)

Although there are many factors that interplay and create multi-dimensional impact on the mental, emotional, physical, and spiritual health of an individual, research findings support that health is maintained by protective lifestyle behaviors. This can include diet and exercise but also reducing stress through such activities such as conscious breathing and meditation. Our respondents told us that poor prenatal *Jing* can be balanced by lifestyle choices.

### BELONGING

Definition: *An inner sense that you matter in the world which can increase or decrease during various times in your life. Belonging is generated in the context of relationships with other people, the environment, and yourself. While belonging can protect against negative context, a lack in the sense of belonging can also lead to poor health outcomes*.

The following quote demonstrates that despite the environment being less than ideal, the relationship between individuals can create a protective factor, leading to a sense of belonging. Additionally, the interviewee offers a snippet into a potential health outcome where both the environment and the relationship with others are solid, perhaps creating another protective layer in the sense of belonging.

> I think that family is one of the most important things. So here’s where we can flip things around. What I’ve seen is that a lot of times in areas that are more depressed, there’s a tighter knit family, group, families living together, grandmothers living in the house supporting each other, and the family status, if we can have a good family status, then that can be very beneficial. (Subject 5: 56:19)

Within the context of a healthcare relationship, a sense of belonging is required for both the patient and the provider to optimize treatment outcomes.

> Being persistent. Showing up, giving time. (Subject 4: 41:14)

Two subcodes of how to promote a feeling of belonging were listening and trust; the above quote highlights aspects that are necessary in order to foster a sense of belonging. When a patient gets bumped from one healthcare provider to the next, it might feel like they are not being heard, as they are often repeating themselves in order for the new provider to understand the background story. Equally, a provider is tasked with staying present and having curiosity in order to cultivate understanding in an effort to create optimal health outcomes. EAM can offer intervention to improve a sense of belonging by nurturing three components at the same time: relationship to self, relationship to others, and the environment.

> There’s three forms of action that we are creating: mind, body, and spirit. And those three forms of action have reactions. If they’re pure or virtuous, it’ll be a more positive situation. For us, some are neutral, and some are negative. So there’s positive, neutral, negative. And I think that it’s important to understand that any one of us can decide in our mind that we are going to be responsible for our thoughts and try to transform the negative thoughts into positive, then that will transform our lives. (Subject 5: 30:43)

The above quote specifically hones in on the self reflection and self awareness components that impact a sense of belonging.

> So, that is interesting that it (belonging) is found throughout many different philosophies. And I think that that comes back to the human being looking into themselves and discovering what is already in them. So it doesn’t matter where you’re from in the world, you’re finding these things are available and the wisdom is within you. (Subject 5: 6:54).

At the heart of it, if someone has an internal relationship to self that is strong and meaningful, this can protect against potentially negative aspects: whether that be from the physical environment or relationships with others, and other factors that can negatively influence one’s relationship with self. If the environment and surrounding relationships are also positive, the individual will possess a sense of belonging that acts as a protective layer for all three pillars to work synergistically. The ability to refine oneself through self discovery allows new information and wisdom to bubble to the surface.

> I would say beyond good food, the ability to socialize with others, the ability to connect with others, the ability to regulate your own anxieties in a social milieu so that you can have friends, so you can be connected to other people. So you can have people be willing to teach you who have things to teach you. Because beyond our family, we have to be able to function socially in the world. And if you don’t have that, life is going to be rough. (Subject 8: 50:00)

Belonging is one of our fundamental codes because it became clear that it is a core foundational piece in which there are several subcodes – family, adult influencer, community, self, and Essence – that could be captured within this code. We wrestled with the concept of which came first: a sense of belonging and its impact on the *Three Treasures (Jing, Qi, Shen*) or vice versa. This robust discussion led us to create a working definition that demonstrated the multi-directional force between a sense of belonging (or lack thereof) and the *Three Treasures.* This definition was based on the outcomes of our research and the association between belonging and health status, most importantly as they are perceived by the patient.

> Truly unconditional love can be a protective [and/or] positive factor. (Subject 7: 42:57).

Belonging can have an impact on an individual’s ability to navigate the world, self-regulate, etc. It can be impacted by others but ultimately boils down to the individual and how they feel and see themselves within the context of the world. This context influences perceptions of lived experiences creating a state of mind and a narrative that will impact how an individual can cope and process experiences. With the fastest growing prevalence of illness being behavioral and/or with a mind-body component, solidifying Belonging as a main code was a startling discovery from our research, as it can be addressed far more holistically than what is currently offered by contemporary BM. EAM is deeply rooted to address the mind, body, and spirit *(Three Treasures)* in every and all treatments. This connection was the final culmination in understanding how the *Three Treasures* cannot be separated and that despite lower *Jing* at birth, how various interventions can act as supportive protections to enable an individual to still thrive by utilizing the other two *Treasures*.

## DISCUSSION

We originally intended to study *Jing,* but our findings quickly demonstrated that the *Three Treasures* are intimately intertwined and *Jing* cannot be isolated as we originally expected. Equally noteworthy was our research highlighting: Measures existing outside the Center for Disease Control (CDC)/WHO’s definitions of DOH that exist within the 10 Key Questions framework. These measures offer novel insight for the advancement of public health, should contemporary medicine be interested.

## BELONGING DISCUSSION

Lack of community or a sense of belonging can be directly correlated to an increased risk for heart disease, high blood pressure, depression, anxiety, suicide, and other comorbidities. The social context or environment is just as important to understand as is the individual patient. In healthcare, a sense of belonging is required for both the patient and the provider to optimize treatment outcomes. In EAM, *health* is seen as a state of ease where the body, mind, spirit *(Three Treasures),* and emotional state are interacting fluidly and sustainably, with *sense of belonging* at the core for wellbeing, and survival. In some instances, as our research clarified, the patient’s internal sense of belonging can have protective effects for their physical, social, emotional, mental, and spiritual wellbeing.

## THREE TREASURES DISCUSSION

The *Three Treasures* are the foundational and key elements in all pattern recognition and therefore we argue that they are in fact the DOH. The *Three Treasures* in EAM (*Jing, Qi and Shen*) are illustrated in the ancient classical texts within the structure of a triangle (Figure 4). Profound modifiability exists, where the *Three Treasures* can compensate for one another in order to create balance and optimal health. For example, if there is discrepancy or deficiency in one “amount” of a treasure, nourishing the other two will help to create and stimulate equilibrium.

**Figure 4:**
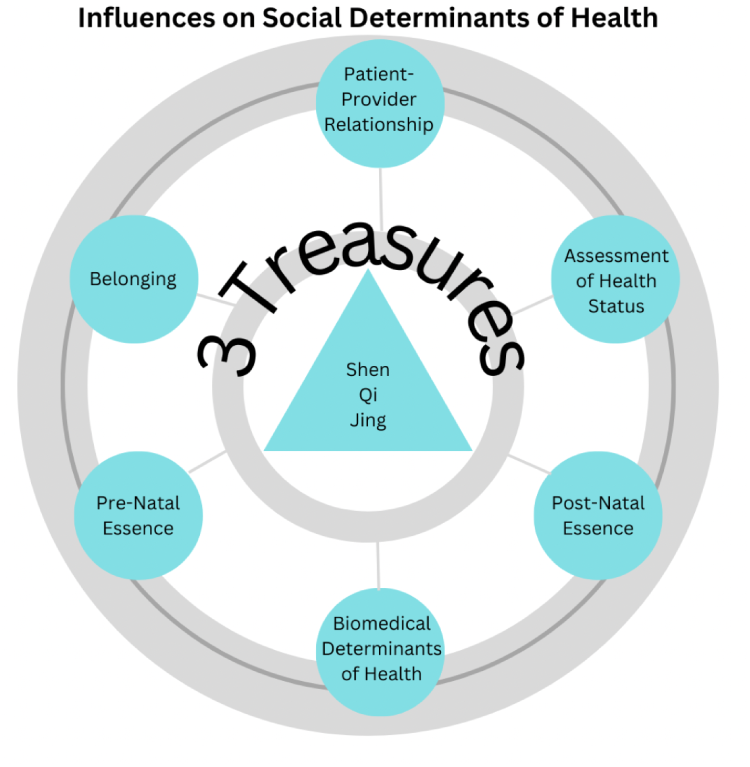
EAM Influence on Social Determinants of Health.

Although our intention was to study only *Jing*, our interviewees and research demonstrates that *Jing* cannot be singularly removed from the *Three Treasures* and examined through a one dimensional framework. Information from the patient is gathered through the 10 Key Questions model, in order to assess the state of the *Three Treasures (*EAM version of DOH) and then come to a diagnosis. Our respondents tell us that through clinical evaluation, relationship building, and an emphasis on developing a compassionate patient-provider relationship that relies on the inner knowing of all parties involved, that the *Three Treasures* can in fact be measured, used to determine health status, and ultimately guide treatment principles to directly impact treatment outcomes.

## PATIENT PROVIDER RELATIONSHIP DISCUSSION

This code is intimately connected to the code, Belonging (see page 13). Listening and Boundaries are two of the sub-codes that emerged from the research.

The patient provider relationship is a focal point of our research. Subcodes such as trust, boundaries, compassion, listening, etc. were discussed as elements that help make up bedside manners. Trust is an integral component of the patient provider relationship and must be given by each involved party. High trust can encourage healthy communication – keeping the patient from getting stuck in their own way of thinking. In certain situations (such as delivering statistics about a health outcome) if there is high trust, a lack of compassion or tactlessness may be overlooked.

A pivot between trust and compassion is listening. Strong listening skills can allow a provider to crucially understand and ‘see’ what might be supportive for the patient. Active listening requires paying attention to not only the verbal but also the non-verbal such as body language, eye contact, etc. Strong listening skills also provide space to inquire when unclear, to reflect back on what is being said – both of which deepen a connection and can build trust and compassion.

A key component of the patient provider relationship is stepping out of the hierarchy that often exists within medicine, which establishes the provider in a higher rank than the patient. In many ways, EAM practitioners approach treatment as a conduit for their patient’s healing – it is ultimately the patient who in fact does their own healing. EAM practitioners can help support the healing discovery: this is important because as healthcare providers, we can only meet a patient where they are– while we can attempt to share our thoughts and offer guidance, it is essentially up to the patient to decide whether or not to take our advice.

The ability to listen, reflect, and ask questions are qualities within a patient provider relationship that can foster trust, intimacy, and a shared reality of healthcare outcomes.

## ASSESSMENT OF HEALTH STATUS DISCUSSION

There are some main components included in a health assessment: the patient provides desired goals or treatment outcomes prior to meeting with their healthcare provider. Additionally, a health assessment can include health history, health goals and outcomes, an evaluation of meeting those health goals and outcomes, diagnosis, etc. Our respondents told us that EAM actively integrates looking at the mind-body-spirit *(Three Treasures)* and offers various modalities to calm the mind, ease the body of pain, and lighten the spirit. In EAM, the diagnostic process is holistic using inspection, palpation, percussion, and auscultation. This includes but is not limited to taking pulses at three different locations on each wrist (six pulse read) and tongue diagnosis. All of this information can be used within the assessment of health, making society the treatment room: everything is diagnostically relevant.

## PRENATAL ESSENCE DISCUSSION

Prenatal Essence captures this concept of energetic potential, as each new life is created and influenced not just by parent 1 and parent 2 coming together, but is also potentially affected by all ancestors. This can account for an outcome looking different than expected, based on how the *Three Treasures* interact with one another.

This “ancestral blueprint” also exists in contemporary BM: the female fetus develops all of the oocytes (eggs) in her lifetime– no oocytes are developed after birth. *Jing* is nurtured and passed down generationally through the bloodline as the ancestral blueprint to influence generations to come. The lifestyle you live now will affect your great great great grandchildren. This principle is supported by evidence based medical research through the understanding of preparing for pregnancy. The health of the mother will be reflected in the growth, birth, and development of the fetus.

Prenatal Essence can also be impacted significantly by trauma, whether it be physical, mental, emotional, spiritual, or environmental trauma. Trauma has significant consequences on the *Jing, Qi*, and *Shen* of individuals and by default, can ripple out into future generations and be measured as DOH. Some examples of current research on trauma offers some options to help the individual manage the aftermath of trauma, but like our research, the most effective therapies have run into various barriers due to lack of funding to conduct further research. Most funding is backed by and set aside for pharmaceutical options (Kolk M.D, B. V. D. 2015). Our research provides an important call-to-action regarding the power each individual has to create a different outcome, if they are able to regulate and practice lifestyle practices that protect the body, mind, spirit *(Three Treasures)*.

## POSTNATAL ESSENCE DISCUSSION

In contrast to prenatal Essence, the “energetic blueprint” of postnatal Essence is influenced by a wider variety of factors, including the addition of weaving a new storyline from the accumulation of ancestors and parents’ *Three Treasures* that create different health outcomes for the individual.

There is a misconception that poor health outcomes might only exist in communities whose mean income is within a certain “lower” margin. However, our respondents provided examples of resilience in lower income groups, demonstrating the equilibrium swing between the *Three Treasures* to create a health better than projected. One respondent gave a clear illustration of how nurturing young minds and lives by teaching music, acting, or participation in other forms of art has allowed people in inner cities to create a different health outcome - the postnatal lifestyle is a constant “dance” of the *Three Treasures* moving towards equilibrium. Another interviewee demonstrated how this is achievable, noting that health outcomes are influenced by multiple lifestyle factors such as nutrition, proper hydration, physical movement, perceived stress levels, consciousness of breath, sleep, meditation, hobbies, and more; parallel factors with the WHO and CDC’s list of DOH. Regardless of socioeconomic status, as our research showed, there is stress of money and time. In many cultures around the world, the term “health” is synonymous with longevity. But health also includes *quality of life*: physical, mental, spiritual, and emotional wellbeing. In short, postnatal Essence is modifiable and many factors influence it.

## DETERMINANTS OF HEALTH (DOH) DISCUSSION

EAM acknowledges that the DOH can pivot between adverse or protective based on an individual’s life context. EAM supplies many ways for the patient and the provider to intervene: patients are encouraged to observe their own health patterns, and work in collaboration with the healthcare provider to identify both patterns of disharmony and possible remedies. The patient is never viewed without their larger social and economic context. With lifestyle choices continually changing the context of whether a determinant of health is aversive or protective, one can see how the *Three Treasures* are constantly seeking equilibrium. And while our research directly pointed out that *Jing* cannot be separated out and analyzed in a one-dimensional manner, by default it also demonstrates that *Jing* is constantly slightly changed as it balances within the *Three Treasures* to optimize health.

## CONCLUSION

Respectful investigation toward understanding EAM systems at the theoretical level can allow for a truly integrated medical system. Although we are familiar with EAM theory, the data surprised us by indicating that *Jing* cannot be considered independently from the *Three Treasures*.

According to the EAM model, health is defined from the position of innate and anticipated longevity through nurturing oneself and cultivating one’s own *Jing, Qi,* and *Shen*. Health is often referred to as a state of ease, synergy, and flow which are not limited to age, gender, or social status but instead ignited and nurtured by a deeper knowing and respect of one’s inner self/essence. Health is a state of being; a verb describing generation that always has direction and potential areas of cultivation. This dynamic definition of health allows for a dynamic view of the DOH, which encourages new areas of thought in the study of health and health promotion.

Currently the fastest growing prevalence are illnesses of behavior or with a mind-body component (e.g. depression), and those that rely on self care. Beyond health and healthcare, we have a growing differential between rich and poor: a gap that will be harder to ignore with a greater acceptance of the negative effects of resulting social DOH. Several research respondents referenced the tension and challenge that exists when isolating aspects of the human experience to trackable data points because this action negates the perspective of whole person health. When a patient is seen as a living organism living in an influential context, it allows for the consideration of many ways to treat the same problem. This flexibility of EAM is a strength, as healthcare truly shifts towards individualized treatment that is comprehensive, and person-centered. There is space in the EAM medical model to treat the patient’s experience; there is always something that the provider can do: it is real-time treating. Additionally, the importance of the patient provider relationship encourages patients to engage in health promoting behaviors. Our work supports the importance of looking at successful ways of organizing society that reduce the negative effects of social differences, and the importance of considering the whole person, with their social, emotional, and subjective states.

## LIMITATIONS

Qualitative research often seeks meaning rather than data to represent a larger population. Our purposive sample of twelve individuals gathered from a wide range of backgrounds, yet future work should include other viewpoints. Our sample size was heavily female-dominated despite outreach to male participants. Additionally, all of our respondents were college educated.

## FUTURE RESEARCH

We set out to address the question of whether lower socioeconomic status would result in less *Jing*. Future collaboration and education among various medical practices, including traditional approaches, would in turn enhance patient assessment/care. Our results suggest that there are many opportunities for the medicines (EAM and BM) to communicate with each other and make a better system of understanding health and patient care. Further research on medical terminology that already exists, overlaps or is completely novel to each medical model is a necessary starting point. Public health would be furthered with the inclusion of *Jing* as a measurement - in many ways, it already is: benchmark for weight, height, scream, etc. at birth.

There are multiple possible studies to explore whether *Jing* at birth shifts with the ‘right’ interventions (e.g. a premature baby in the last percentile at birth that is found to be above the 50% percentile at their one-year check up) as postnatal *Jing* is formed. Another possible study would follow babies of similar benchmarks at birth through their childhood and into their teenage years.

Our research findings can be extrapolated and further research should be conducted in order to prove or disprove the following:

i. The importance of assessing an individual as a whole person: mind, body, and spirit *(Three Treasures)*.
ii. Current social epidemics highlight the current gaps in serving the public because the offered solution does not address mind, body, and spirit *(Three Treasures)* together. Aspects of public health solutions have failed due to the attempt of addressing the *Three Treasures* as an individual one-dimensional aspect, just as we approached the research of looking at *Jing.* Possible examples include homelessness, drug addiction, etc. where the few public health solutions could in fact address/have a mind, body, spirit solution (*Three Treasures)*.
iii. In the EAM model, the *Three Treasures* are the DOH. With continued efforts for collaborative and integrative care, explore how the DOH as defined by WHO and CDC can be in collaboration with the EAM model for the advancement of future integrative healthcare?

## Supporting information

Full Supplemental Document

