## Supplementary material for "Investigating How East Asian Medical and Biomedical Systems Can Communicate: A Qualitative Exploration of How East Asian Medicine Concepts Can be Used in Modern Health Research": Full Supplemental Document

#### Appendix and Table Index

**Appendix 1:** Research Authors Biographies

**Appendix 2:** De-identified Interviewee's Professional Titles and Credentials

**Appendix 3** Consent to Participate

**Appendix 4:** Interview Research Questions

**Appendix 5:** The "10" Key Questions

**Appendix 6:** Figures Key

---

**Table 1:** Terms and Definitions used in East Asian Medicine

**Table 2**: Final Codes, Definitions, and Examples

**Table 3:** Acronym Key

### APPENDIX 1 : Research Authors Biographies

RESEARCHERS

- Rachel Burack, an alumna of the New England School of Acupuncture, earned a dual degree in Acupuncture and Chinese Herbal Medicine. Her academic and clinical excellence was recognized through the prestigious Compassionate Healer Award. In her patient-centered private practice, Rachel specializes in family, pediatric, and psychosocial medicine, focusing on educating and empowering her patients. Her career, spanning over a decade, is marked by a rich diversity of experiences. These include advanced studies in yoga, committed service with AmeriCorps, and extensive travel, all of which have contributed to her broadened worldview and empathetic approach in both her professional and personal life. Rachel's deep-seated passion for holistic medicine goes beyond her professional endeavors, reflecting her personal dedication to the field's advancement.

- Lisa Conboy has been researching Complementary and Alternative Medicine using quantitative and qualitative research methodologies for over 30 years; 20 of which at the New England School of Acupuncture. She is an Instructor at Beth Israel Deaconess Medical Center, Harvard Medical School, and the chair of the Research Committee of the American Society of Acupuncturists.

- Maria Engels received her Master of Acupuncture from the New England School of Acupuncture and was a recipient of the President's Commitment to Diversity Award, the New England School of Acupuncture Service Award, the Inclusive Excellence Award, and was inducted into the Honor Society of Phi Kappa Phi in 2020. Her prior work experience as a nonviolence educator combined with her life experiences as an international adoptee within a diverse family background influenced her approach to research by cultivating a lens of curiosity towards epistemology.

- JoHannah Macbeth is a graduate of the New England School of Acupuncture and holds a dual degree in Acupuncture and Traditional Herbal Medicine, with specialization in Japanese and Orthopedic Styles. During her academic term, JoHannah piloted an Integrative Clinical internship program at the prestigious Cleveland Clinic for which she earned the Tsai Award. Currently, she treats patients at clinics in both Massachusetts and Rhode Island, including the Community Wellness Clinic at MIT in Cambridge, MA. JoHannah also maintains a private practice in Rhode Island for Integrative and Therapeutic Bodywork. In addition to her clinical work, several previous experiences offered JoHannah distinctive insights for clinical and cultural references during this project, including teaching and performing as a classically trained musician, adjunct teaching at the Community College of Rhode Island, and serving on rehabilitation teams in underserved communities across the globe."

### APPENDIX 2: De-identified Interviewee's Professional Titles and Credentials

For privacy, the interviewee's name have been concealed and they will be identified by their professional titles and distinctions.

| **Name:** | **Title/Credentials:** |
| --- | --- |
| Subject 1 : | LAc |
| Subject 2: | Master Face Reader, Lotus Institute Founder |
| Subject 3: | Social Justice Educator, Nonprofit |
| Subject 4: | CEO, Ellis Early Learning |
| Subject 5: | Professor of Mindfulness Studies, Yoga instructor |
| Subject 6: | LAc |
| Subject 7: | Professor, Academic Advising, Health and Wellness Coach |
| Subject 8: | LAc, Podcast Host |
| Subject 9: | LAc, DAOM, Clinical Researcher |
| Subject 10: | Family Systems Coach, LCSWA |
| Subject 11: | UC Berkeley Lab Researcher in Bioengineering |
| Subject 12: | LAc, DAOM |

#### APPENDIX 3: Consent to Participate

**TITLE**
Investigating How East Asian Medical and Biomedical Systems Can Communicate: Jing Poverty

**PURPOSE**
You are being invited to participate in a research study. Research is a way of gaining new knowledge. The purpose of this research is to learn about how experts like you view the relationship between generational low income/ socioeconomic status and inherited jing (a Chinese Medicine concept similar to biological resilience)

**WHO MAY PARTICIPATE**
In order to participate in this research, you must be a professional in the science of acupuncture and/or medical science, and be at least 18 years of age. If you are not a professional in the science of acupuncture and/or medical science, or if you are not at least 18 years of age, you should not participate. MCPHS students and faculty may not participate in this study.

**PROCEDURES**
Participation involves a one-on-one interview over phone or Zoom. The interview will take about an hour. The interviewer will ask your views on health, genetics, and socioeconomic status. You will also be asked to read a brief description of the topic prior to the interview. The reading should take only a few minutes, so the total time for participating will be approximately one hour and ten minutes. With your consent, we will record the interview to assist with later analysis.

**PARTICIPATION IS VOLUNTARY**
You do not need to participate in this research. There is no penalty for deciding not to participate.

**RISKS AND BENEFITS**
This research is not designed to provide direct benefit to participants. The primary benefit of this study will be the opportunity to contribute to knowledge about our understanding of the relationship between jing and poverty.

The only known risk of participating is a small chance that someone outside the research team would be able to see the data you provide and will know that you were the one who provided it. We will take reasonable precautions to avoid this.

**COMPENSATION**
You will not be compensated for your participation.

**CONFIDENTIALITY**
The researchers will be able to see (if you use a webcam) and hear you during the interview. However, we will not use identifying information when we analyze and prepare the results of our research for publication or presentation. The researchers will keep the identity of participants confidential. Data will be stored in a secure electronic device that will be password-protected and accessible only to the researchers. At the conclusion of the study, the data will be securely transferred to the MCPHS University for storage, and after five years, the data will be destroyed.

**CONTACT**
If you have questions about this research, please contact the Principal Investigator of the research:

Lisa Conboy ScD. 617 718 1917.

If you have questions about your rights as a participant in research, please contact the Chair of the Institutional Review Board (IRB) at MCPHS University:

Kenneth A. Richman, PhD
kenneth[DOT]richman[AT]mcphs[DOT]edu.

**SPOKEN CONSENT**
Before the research portion of the interview begins, the interviewer will ask whether you have questions about this document or about the research. You will be asked to say out loud whether you agree to have your answers recorded and used for research.

### APPENDIX 4: Interview Research Questions

These questions were presented visually in a PDF presentation on the screen during each interview.

| QUESTION | QUESTION TEXT |
|---|---|
| Question 1A: | *“As we try to understand more about how the concept of Jing can be used in modern health research, is it necessary to understand similar concepts now in use?”* |
| Question 1B: | *“Are there similar concepts in your science? How are they different or similar to Jing?”* |
| Question 2: | *“We are studying the potential relationships between socioeconomic status and genetics, including epigenetics. Do you think that socioeconomic status is related to genetics and/ or epigenetics?”* |
| Question 3: | *“What methods do you use to identify the state of health in your patients or subjects related to genetics/ epigenetics?”* |
| Question 4A: | *“What economic/ socio-psychological factors might negatively impact a patient’s or subject’s epigenetic presentation?”* |
| Question 4B: | *“How might such processes be linked to trends in health status that we see generation to generation?”* |
| Question 5A: | *“What economic/ socio-psychological factors might positively impact a patient’s or subject’s epigenetic presentation?”* |
| Question 5B: | *“How might such processes be linked to trends in health status that we see from generation to generation? “* |
| Question 6A: | *“Is it necessary to build a relationship with your patient or subject to do your work?”* |
| Question 6B: | *“In your experience, what is the most impactful way to build a relationship with your patient or subject?”* |
| Question 6C: | *“Is it important that the patient or subject is comfortable with the relationship? How would you define “bedside manner?”* |
| Question 7: | *“Did you feel a spark to share something additional? Please share any additional information relevant to this topic and research you feel is pertinent to be considered.”* |

### APPENDIX 5: The “10” Key Questions

Traditional diagnostic questions for the practitioner of East Asian Medicine.

| Category | Questions |
|---|---|
| **PAIN** | Assess Pain using OPQRST:<br>● Onset: When? Sudden or progressive?<br>● Provocative/ Palliative factors: What makes it better? Worse? Better or worse w/ pressure? Better or worse with movement? Better or worse w/ heat or cold?<br>● Quality: sharp/dull, aching/stabbing, crushing, burning?<br>● Radiation: Does the pain travel?<br>● Severity: on a scale of 1-10<br>● Time: continuous/ sporadic? day/night? Has it occurred before? |
| **FOOD and TASTE** | ● Does eating make it better or worse?<br>● Preferred type of food? Hot or cold?<br>● Any taste in the mouth?<br>● Nausea/ vomiting? What causes it? Vomit description |
| **STOOLS and URINE** | ● Bowel movements: Daily? How often? Dry/sticky/ loose/watery? Smell? Formed or Unformed?<br>○ Constipation/ diarrhea or both? Any recent changes?<br>○ What color is the stool? Ever noticed any blood?<br>● Urination: Any problems stopping or starting? Any pain? Color? Frequency? |
| **THIRST and DRINK** | ● Feeling thirsty? Dry mouth? No thirst?<br>● How much water per day<br>● Preference for hot, cold or room temperature drinks? |
| **HEAD (+FACE) and BODY (+ LIMB)** | ● Headaches? Dizziness? Whole body pain? Joint pain? Backaches? Numbness/ tingling?<br>● Use OPQRST to describe |
| **CHEST and ABDOMEN** | ● Chest pain? Epigastric pain? Lower abdominal pain? Hypogastric pain? Abdominal distention?<br>● Use OPQRST to describe pain<br>● Worse or better w/ bowel movements or food? |
| **SLEEP** | ● Hours of sleep each night? Regular sleep schedule?<br>● Does it feel like enough? Tired during the day?<br>● Any recent changes in sleep?<br>● Difficulty falling asleep? Staying asleep? Waking up?<br>● Dreams/ nightmares? |
| **SWEATING:** | ● Amount: (too much/ not at all)?<br>● What activities cause sweat? What time of day/ night? |

| | |
|---|---|
| | • Quality of sweat? (Oily, watery, cold, sticky) |
| **EARS and EYES** | • Ears: Tinnitus? Onset? Pressure? Describe noise? Pitch?<br> ○ Deafness: Sudden or gradual<br> ○ Itching: Exudate?<br>• Eyes: Pain? Dryness? Tearing? Distension? Itchy? Blurred vision? Decreased vision? Onset? Sudden or gradual? |
| **CHILLS and FEVER** | • Any chills/ fever/ both?<br>• Alternating chills and fever?<br>• Better/worse with heat, cold or drafts? |
| **ENERGY LEVELS** | • Scale of 1-10 or Low/Medium/High<br>• What time of day is the highest? Lowest?<br>• Better/ worse after meals?<br>• Better/worse after rest?<br>• Energy level reasonable based on daily lifestyle/ schedule? Age? |
| **EMOTIONS** | • How do you feel about that?<br>• Ever feel sad? Depressed? Angry? Worried? Scared? |
| **SEXUAL SYMPTOMS** | • Is the condition aggravated by sexual activity?<br>• Excessive fatigue or headache after sexual activity?<br>• Any sexual difficulties to mention? |
| **WOMEN's SYMPTOMS** | • Menarche? Regular menstrual period?<br>• Last menstrual period? How many days of beedling? Day between? Total cycle lengthy?<br>• Any PMS symptoms? Feel better before or after period?<br>• Color of blood? Amount? Clotting?<br>• Pain/ cramping? Digestive changes? Headaches? Emotional changes?<br>• Menopause? Symptoms? |
| **CHILDREN'S SYMPTOMS** | • Patient input is key<br>• Listen to and/or observe the child as much as possible<br>• Difficulties during the pregnancy/ birth/ breastfeeding?<br>• Childhood disease? Immunizations?<br>• Digestive symptoms? Respiratory? Earache? Sleep?<br>• Development: Standing? Teeth? Hair? Walking? Speech? |

Citation:

Kim H. (H. B. (2015). *Minibook of oriental medicine* (Third). AcupunctureMedia.com. pg. 41

### Appendix 6: Figures

Figures are listed below in the order that they appear in the published paper.

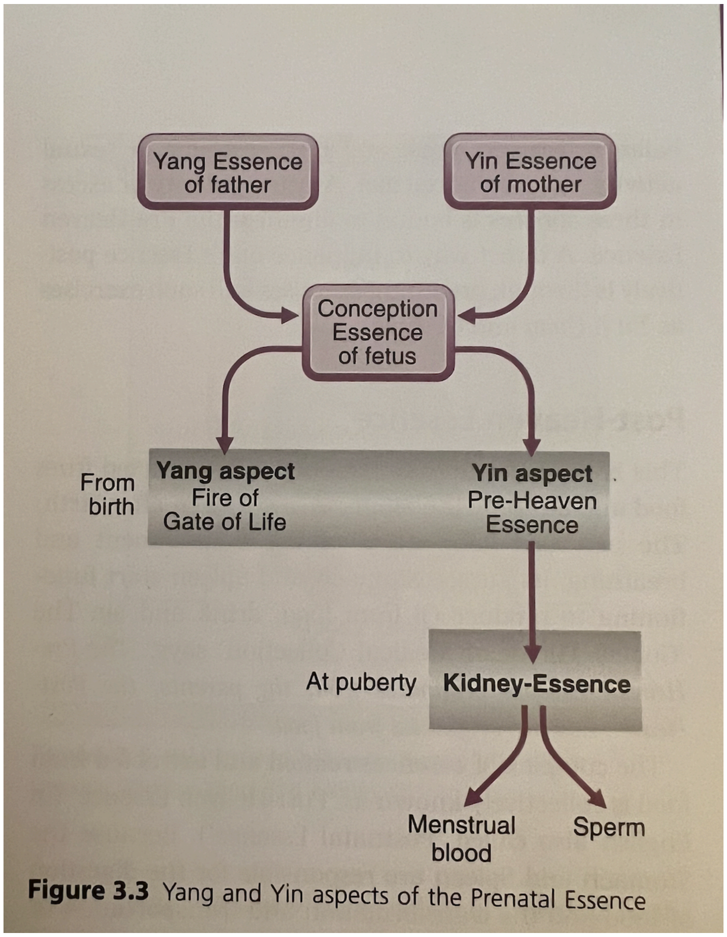


**Figure 3.3** Yang and Yin aspects of the Prenatal Essence

**Figure: 1**
**Description:** “Yang and Yin Aspects of the Prenatal Essence”
**Source:** Maciocia, G. (2005). *The foundations of Chinese medicine: A comprehensive text for acupuncturists and herbalists*. Elsevier Churchill Livingstone. pg 47

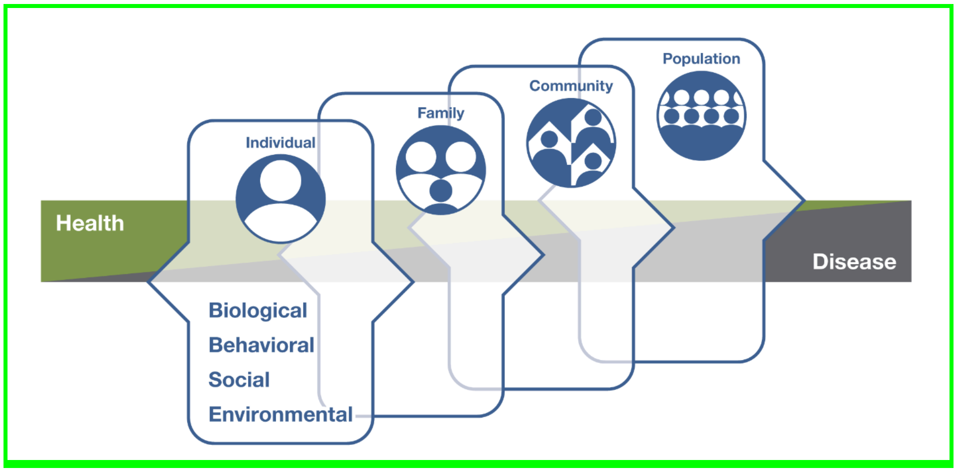


**Figure 2:**
**Description:** Factors Influencing Health and Disease.
**Source:** https://www.nccih.nih.gov/health/whole-person-health-what-you-need-to-know

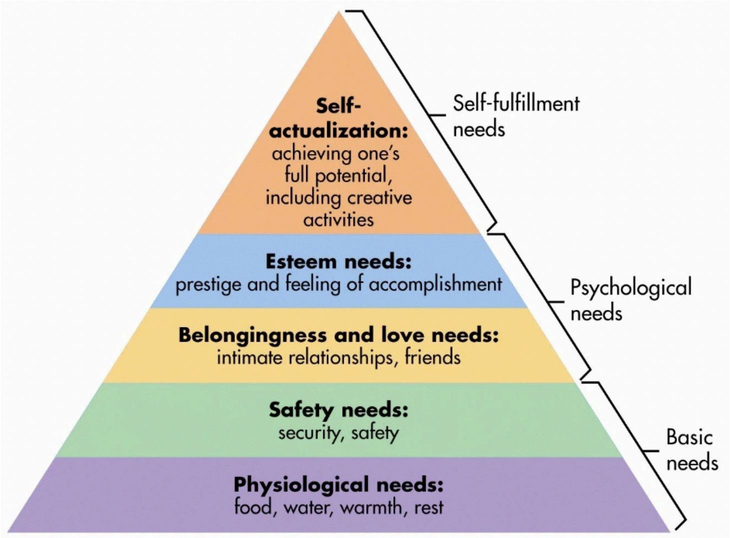


**Figure 3:**
**Description:** "Figure 2.0 Maslow Hierarchy of Human Needs: A Humanistic Psychology Approach of Abraham Maslow
**Source:** Kawanua International Journal of Multicultural Studies, Vol. 3, No. 2, December 2022, p. 30~35, ISSN: 2797-5460, E-ISSN: 2797-359X, DOI: 10.30984/KIJMS.v3i2.282

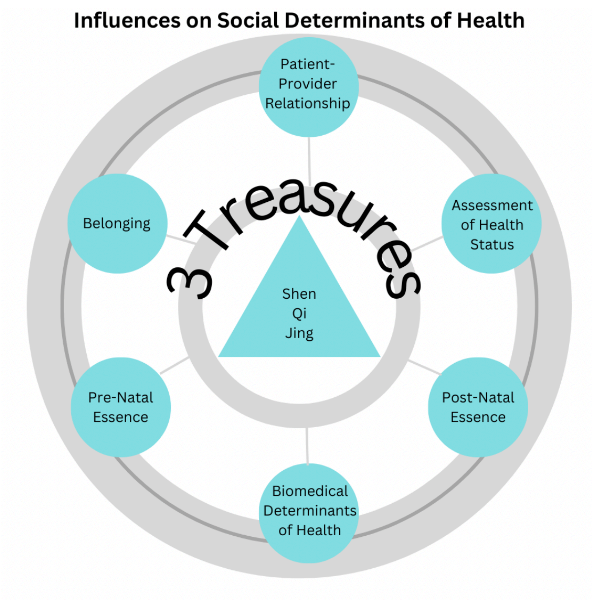


**Figure 4:**
**Description:** East Asian Medicine Influence on Social Determinants of Health
**Source:** Made by Rachel Burack

### Table 1

**Terms and Definitions used in East Asian Medicine**

| TERM | DEFINITION |
|---|---|
| **Belonging** | *"A sense of belonging—the subjective feeling of deep connection with social groups, physical places, and individual and collective experiences—is a fundamental human need that predicts numerous mental, physical, social, economic, and behavioral outcomes."* [1] |
| **Biomedicine** | *"Medicine based on the application of the principles of the natural sciences and especially biology and biochemistry."* [2] |
| **Determinants of Health** | *" The determinants of health include: the social and economic environment, the physical environment, and. the person's individual characteristics and behaviors."* [3] |
| **East Asian Medicine** | *" East Asian Medicine (EAM), of which Acupuncture is a part, encompasses an entire system of medicine that has many branches and traditions. Although there is debate in the EAM community about the best name for this system of medicine, "East Asian Medicine" is an inclusive term for the medicine born from East Asia. Can be used synonymous with Traditional Chinese Medicine, Acupuncture."* *(In text quotation)* |
| **Epigenetics** | *"The study of heritable changes in gene function that do not involve changes in DNA sequence."* [4] |
| **Genetics** | *"1. A branch of biology that deals with the heredity and variation of organisms 2. The genetic makeup and phenomena of an organism, type, group, or condition."* [5] |
| **Health Disparities** | *"Health disparities are preventable differences in the burden of disease, injury, violence, or opportunities to achieve optimal health…."* [6] |
| **Health Status** | The patient's state of health. EAM and Biomedicine have somewhat different ways of measuring this. |

| | |
|---|---|
| **Jing (Essence)** | *"Jing is usually translated as 'Essence'. The Chinese character for 'jing' is composed of the character for 'rice' on the left and that for 'clear, refined' on the right. Thus the character for 'Essence' gives the idea of something derived from a process of refinement or distillation: it is a distilled, refined essence, extracted from some coarser basis. This process of extraction of a refined essence from a larger, coarser substance implies that the Essence is a rather precious substance to be cherished and guarded."* 7 |
| **Microsystems (of EAM)** | *"TCM uses the body's microsystems to treat the whole person effectively, mirroring a holographic perspective of each part reflecting the whole. This can be seen in various parts of the body reflected in multiple areas within the face, eyes, ears, hands, and feet. On a therapeutic level, TCM addresses these through acupuncture point manipulation. As an example: auricular acupuncture, hand and foot reflexology, etc. "* 8 |
| **Patient- Provider Relationship**<br>Alternatively called patient-clinician, patient-physician… | "*The dynamic therapeutic relationship between the patient and health care/ service provider, often referred to as the practitioner.*" 9 |
| **Prenatal essence**<br>*Energetic blueprint* | *" ..also translates as Congenital Essence, is inherited from the parents. In fact the fusion of this parental Essence is conception. Each person's Prenatal Essence is unique and will determine his or her particular growth patterns. The quantity and quality of the Prenatal Essence is fixed at birth and, together with Original Qi, determines an individual's basic makeup and constitution."*<br><br>Energetic Blueprint: Prenatal essence is the foundational idea behind the concept of *"energetic blueprint."* 10 |
| **Postnatal essence** | *" … is the second source and aspect of Essence. It is derived from the purified part of ingested food and continuous physical, emotional, and mental stimulation from a person's environment. The post Essence allows for modification of prenatal Essence. Together they compose the overall Essence of the person."* 11 |

| | |
|---|---|
| **Qi** | *" Chinese medicine sees the working of the body and mind as a result of the interaction of certain vital substances. These substances manifest in varying degrees of 'substantiality', so that some of them are very rarefied and some totally non-material. All together, they constitute the ancient Chinese view of the body-mind. In Chinese philosophy and medicine, the body and the mind are not seen as a mechanism (however complex) but as a vortex of Qi in its various manifestations of interaction with each other to form an organism. The body and the mind are nothing but forms of Qi. At the basis of all is Qi: all the other vital substances are but manifestations of Qi in varying degrees of materiality, ranging front the completely material, such as Body Fluids, to the totally immaterial, such as the Mind."* 12 |
| **Social Disparities**<br>Determinants of health<br>Social determinants of health | Determinants of Health According to WHO as of 2013:<br><br>*"Many factors combine together to affect the health of individuals and communities. Whether people are healthy or not, is determined by their circumstances and environment. To a large extent, factors such as where we live, the state of our environment, genetics, our income and education level, and our relationships with friends and family all have considerable impacts on health, whereas the more commonly considered factors such as access and use of health care services often have less of an impact."* 13<br><br>Social Determinants of Health :<br><br>*"Social determinants of health (SDOH) are the conditions in the environments where people are born, live, learn, work, play, worship, and age that affect a wide range of health, functioning, and quality-of-life outcomes and risks."* 14 |
| **Three Treasures**<br>(Jing, Qi, Shen) | *" The Essence and Qi are also considered to be the material foundation of the Mind (Shen). Essence, Qi and Mind (Jing, Qi, Shen) are the three fundamental physical and psychic substances of a human being. For this reason, they are called the Three Treasures. Essence, Qi, and Mind also represent three different states of condensation of Qi, the Essence being the densest, Qi being rarefied, and the Mind being the most subtle and immaterial. According to Chinese medicine, Essence and Qi are the essential foundation of the Mind. If Essence and Qi are healthy and flourishing, the Mind will be happy and this will lead to a healthy and happy life. If Essence and Qi are both depleted, then the Mind necessarily will suffer."* 15 |

| | |
|---|---|
| **Yi** | *" Consciousness of Potentials, or Yi: Yi is sometimes translated as thought, consciousness, or intention, and, in this book, it is translated by the cumbersome but more accurate phrase "Consciousness of Potentials." Operationally, it is the Spirit that is responsible for considering, deliberating, and deciding on what is likely, possible, or conceivable. It is responsible for discerning various directions and perspectives. The Consciousness of Potentials is ultimately responsible for allurement, vision, motivation, and creativity."* 16 |
| **Shen**<br>(Spirit) | *" The Mind (Shen) is one of the Vital Substances of the body. As we have seen above, the Vital Substances take shape in different degrees of aggregation of Qi: the Mind is the most subtle and non-material type of Qi. The word Shen is often translates as 'Spirit' in the Western acupuncture books and schools; but I prefer to translate Shen as 'Mind' since I believe that what we would call 'Spirit' in the West is the complex of all fiver mental-spiritual aspects of a human being: i.e. the Ethereal Soul (Hun) pertaining to the Liver: the Corporeal Soul (Po) pertaining to the Lungs: the Intellect (Yi) pertaining to the Spleen: the Will-power (Zhi) pertaining to the Kidneys: and the Mind (Shen) itself"* 17 |
| **Whole Person Health**<br>Synthesis<br>Analysis | *"Whole person health involves looking at the whole person—not just separate organs or body systems—and considering multiple factors that promote either health or disease. It means helping and empowering individuals, families, communities, and populations to improve their health in multiple interconnected biological, behavioral, social, and environmental areas. Instead of treating a specific disease, whole person health focuses on restoring health, promoting resilience, and preventing diseases across a lifespan."* 18 |

9. Rekindling Joy in Medicine Through Thoughtful Communication: A Practical Guide
Tara Sanft and Eric Winer. American Society of Clinical Oncology Educational Book 2023 :43

10. Kaptchuk, T. J. (2000). The Web That Has No Weaver (3rd ed., pp. 55-56). McGraw Hill.

11. Kaptchuk, T. J. (2000). The Web That Has No Weaver (3rd ed., p. 56). McGraw Hill.

12. Maciocia , G. (2015). The Foundation of Chinese Medicine (25th ed., p. 43). Elsevierl.

13. *Determinants of health*. (n.d.). https://www.who.int/news-room/questions-and-answers/item/determinants-of-health

14. *Social Determinants of Health - Healthy People 2030 | Health.gov*. (n.d.). https://health.gov/healthypeople/priority-areas/social-determinants-health

15. Maciocia , G. (2015). The Foundation of Chinese Medicine (25th ed., p. 50). Elsevier.

16. Kaptchuk, T. J. (2000). The Web That Has No Weaver (3rd ed., pp. 59-60). McGraw Hill.

17. Maciocia , G. (2015). The Foundation of Chinese Medicine (25th ed., p. 70). Elsevier.

18. *Whole Person Health: What You Need To Know*. (n.d.). NCCIH. https://www.nccih.nih.gov/health/whole-person-health-what-you-need-to-know

#### Table 2: Final Codes, Definitions, and Examples

| Code Name | Quotes from Data |
|---|---|
| **BELONGING**<br><br>**Definition:** An inner sense that you matter in the world which can occur on a spectrum during various times in your life. Belonging is generated in the context of relationships and can be positively or negatively related to health status.<br><br>**Related Codes/Themes:**<br>● Family<br>● Adult Influencer<br>● Community<br>● Self<br>● Essence (Western)* | *"I think that family is one of the most important things. So here's where we can flip things around. What I've seen is that a lot of times in areas that are more depressed, there's a tighter knit family, group, families living together, grandmothers living in the house supporting each other, and the family status, if we can have a good family status, then that can be very beneficial. It's not always the case, obviously, there's a lot of people, there's many children from different places who don't have parents that are living with them and don't have fathers living with them."* (Subject 5: 56:19)<br><br>*"Terms of families, yes, I have done this, where, you know, you treat grandma and all of sudden, everybody's much better in the household because she's not as irritable." "She's not demanding, she's calmer. She's nicer. She, you know, she doesn't make you feel as inadequate as she was making you feel. Yeah, she's more appreciative. Yeah, I have one gentleman who, who was able to say he loved .... to his daughter. And, you know, like to just be able to open up and be able to say I love you to his daughter was like, so healing to the whole family. Luckily, he because he was dead within like, I think a year or 18 months. And you know, and if, if he had never said that, that would have never healed."* (Subject 12: 37:10 + 37:43)<br><br>*"You know, it really, really depends on what someone is willing to look at. And if they have a family that helps them recognize these, these are things that you went through these the ways that you deal with these things, these things make a change in health status, there's a really important thing that epigenetics So, you know, I actually see that they definitely can be linked to health status. But I find that it's even more likely that epigenetics will actually help people open up from where they've been. So if they have something that's kept them really stuck, when they open up, they really can see something very, very different. And that changes the pattern. So I see something really different happening."* (Subject 2: 42:39)<br><br>*"[Maciocia Image: It becomes like a family tree like it would be the same image for the mother and then her mother, the mother's ancestors in the same image for the father's ancestors, it'd be like this big family tree coming down"* (Subject 6: 2:14)<br><br>*"Truly unconditional love can be a protective/positive factor." (Subject 7: 42:57)*<br><br>*"I would say beyond good food, the ability to socialize with others, the ability to connect with others, the ability to regulate your own anxieties in a social mill, so that you can have friends, so you can be connected to other people. So you can have people be willing to teach you who have things to teach you. Because beyond our family, we have to be able to function socially in the world. And if you don't have that, life is going to be rough."* (Subject 8: 50:00) |

| **3 TREASURES**<br><br>**Definition:** A representation of three factors referenced in EAM as Jing, Qi, and Shen. The unique expression of an individual's core nature which allows the determination of health to be made, influenced by pre-natal (genetics) and post-natal (epi-genetics) factors.<br><br>**Related Codes/Themes:**<br>● Qi<br>● Jing<br>● Shen<br>● Compassion<br>● Happiness | *"The Jing like nourishes or fortifies the body and the Qi will mobilize it. But the Shen engages the body in the world."* (Subject 12: 41:10)<br><br>*"[Participating in the arts] changed my life. Or epigenetically changed me."* (Subject 12: 41:56)<br><br>*"So you have to be, your Jing has to be solid, your Qi has to be flowing and your Shen has to be engaged. So you can hear, see and take in, you have the information you need but not merge. So you have to have good boundaries. So your egg has to be your energetic egg, has to be complete, but not not barriers. You have to let the ship in, you have to let the information in, which comes with this, you know, you're not letting the Jing in right, you're not having sex with your patient, I hope. that and you're not letting so much, you know, you're not letting their Qi in because their aches and pains, you don't want to take it. Just like their relationship to it. Their Shen is really what you're kind of engaging to get the information about the rest, you're touching. So you're using your Qi to feel. So you're getting that but you're not embodying it. It's very, it's very subtle, because you need to have a call. It's co creative, but you need to have your boundaries."* (Subject 12: 47:09)<br><br>*"So whether we're untouchable, or a king, happiness is going to be what dictates and can we have the mindset to try and transform the mind. So although socio economic status, I think makes a difference. And it could be more challenging for us based on what we're born into. It doesn't dictate for us that there is a master plan that we can't get out of, it's not destined that this life is going to be miserable because of our socio economic status, and that our health will be miserable, but that we can overcome."* (Subject 5: 39:56)<br><br>*"Jing, Qi, and Shen, as being something unusual that can be used now, in a way that's very authentic, classical Chinese medicine."* (Subject 2: 3:57)<br><br>*"So the thing is, I don't I don't find that most people are missing their Jing. I don't think they're missing their epigenetics. I don't think they're missing their genetics. I mean, I just, I think that's really easy. I think what's hard is for people to remain constant. So the Shen becomes very much of an issue. And so of course Qi, which is really important, is something that's really really critical, but it's only really helpful if someone's Shen is present. So I tend to focus a bit more in Shen"* (Subject 2: 34:02)<br><br>*"Epigenetics influenced by timing and mental outlook ."*(Subject 2: 40:14)<br><br>*"I actually don't share as much about Qi as you might think, even though that's what Chinese Medicine is about. It's not that its not important, it's very important, but they do know as many things you can do to enhance Qi. I also know that Jing is extremely important. And I think about something I focused on a lot. Because people don't know enough about how to manage their Jing. But I think the most important thing about longevity has to do with their Shen. And if we can find ways that help their Shen, with the big S .. H.. E.. N... we can find ways for that Shen to be to be recognized and helped. And, and really what's the word I want to say that really helped to be seen, I think we're gonna help people live a long, long time. So I really, I think that that Jing is really, really important. I think that Shen is really really important. I think Qi's really important. But of all* |
|---|---|

| | |
|---|---|
| | *those, I'd say, Shen is most important, if you can get to it. If you can't get to it, you have to go through Jing to get it. And so that's something to be aware of. So I'm very .. whats the word I want to say ... I think I'm very clear that it's important for... what's the words I want to use here .... It's important for Shen to have a way of being revealed. that isn't so.... what's the words I want to say?..... Isn't so automatic. Every person is different, when it comes to like what it is about you and your Shen that you can bring through."* (Subject 2: 57:19)<br><br>*"So it's, yes, it's Qi and blood and Jing. The three the three treasures, I think, I think to disregard the potential, the influence of history and the potential reality of future is, in my mind, a little short sighted."*<br>*(Subject 6:12:48)*<br><br>*"Jing is primarily associated with water. It's not the only thing that's associated there, but it is something that is associated. And I do look at it, you know, through the water element. But there's more to it than that. So it's not just water, it's just that that's where the kidneys kind of come from. Okay, well hold on to this, we'll, we'll do something with it. So you can actually hold it in the body. So there's much more to it than that. But I do want to say that there are aspects of, of Jing that are more than just the water element. It's not just water, but water helps to hold it, water helps to put it in a place."* (Subject 2: 20:22)<br><br>*"Pre-heaven essence captures the idea of energetic potential – this concept does not necessarily get captured that way within Western medicine.* (Subject 7: 12:25)<br><br>*"Epigenetics is the most important factor that is affecting your health."* (Subject 11: 6:05)<br><br>*" Jing is qi, and qi is qi. So if you're living a balanced life, by definition, you are not depleting it.* (Subject 1: 1:04:05) |

| **PATIENT-PROVIDER RELATIONSHIP**<br><br>**Definition:**<br>The relationship between patient and provider must be professional and intentionally therapeutic. Patients are seen as whole persons and health-related goals are created in a collaborative and mutual manner to optimize outcomes.<br><br>**Related Codes/Themes:**<br>● Provider<br>● Patient<br>● Wisdom/Intuition/Yi<br>● Eyes<br>● Bedside Manner<br>● Boundaries<br>● Body Language<br>● Listening<br>● Trust<br>● Compassion | *"When we meet with someone. really listen, because that doesn't happen much anymore. We're not really listening. What's happening is we're so how are you doing? Uh huh. And we've got other things going on. And not instead of actually sitting in front of the person and saying, What's going on, and then not telling them what they're, what they're going through? Because so often, we want to interject and tell someone about what they're experiencing and how they should be experiencing things. And instead, I think, if we could take a step back, and just listen, that would be huge to building the relationship in developing an understanding of this patient or subject that we're working with."* (Subject 5: 1:04:18)<br><br>*"The great doctors, look at the tongue, feel the pulse, ask a couple of those really important questions, and they got it. Right. You guys know what I'm talking about? Right? Yeah. I'm terrible at the that. I've never been good at it. I have always had to wait. And listen, until a patient said something and I would go., "ahhh" This is something that they say, usually they're actually telling me what they need, I have to wait until a patient tells me what they actually need."* (Subject 8: 59:30)<br><br>*"[family practice] Okay, I always say more ears in the room, we can hear. And then I can hear your voices because everyone adds something to the conversation."* (Subject 6: 28:48)<br><br>*"Relationship is commensurate with the work that they're actually there to do."* (Subject 8: 53:18)<br><br>*"Respecting one's boundaries [is the most important factor in building a relationship]."* (Subject 9: 41:35)<br><br>*"Healing is a co -creative process. That's the first. The only thing I can say is healing is a co-creative process."* (Subject 12: 44:56)<br><br>*"It's co creative, like, they change me, I change them, we have an interaction, I don't go in and have, I may have an idea what I'm going to do, I certainly should have an idea what I'm going to do, but then there's, they come in, and it's a different day. And I, you know, you have to be present. So it's very necessary to keep yourself healthy, to do your Qi gong to keep yourself clean. So you're not taking on stuff. Very important. Take on not to take on stuff, right? If you're building a relationship, but it's not this, when I say it's co creative, it's not this mutual of I give you give me, it's like, I can't take you. But I can intellectually circulate it right. So I need to be present. My spirit, my Shen has to be there.* (Subject 12: 45:06)<br><br>*"A large portion of the actual treatment and the success of a treatment is listening to a person's story. You don't need to comment on the story. But you need to listen to it. You need to keep boundaries around the story, you need to keep them tight, whatever your comfort level is, like, like, listen to people, and you need to listen to them from your first encounter, if it's a phone call on the phone, you've got to really listen and see where they're coming from. People want to tell you their stories, and you need to listen. We can be really powerful mirrors of people's lives, to help them navigate and make change."* (Subject 1: 26:17) |
|---|---|

| | |
|---|---|
| **ASSESSMENT OF HEALTH STATUS**<br><br>**Definition:**<br><br>The collaborative process between the patient and practitioner where the practitioner gathers information using all of their five senses to integrate information with the foundational framework of the traditional 10 Key Questions* viewing the patient holistically; mentally, emotionally, physically, and spiritually.<br><br>**Related Codes/Themes:**<br>● Listening<br>● Observation<br>● Western Science/Biomedicine<br>● Eastern Science<br>● Inner Science<br>● Individualized Health Care | *"Body language is huge, to actually be open versus closed. And if we are open and have an open heart with the patients, I think they will feel that they have our attention and respect and understanding and treat them."* (Subject 5: 1:05:45)<br><br>*"I think we're a very busy society, constantly on to the next thing without giving time. So when it comes to bedside manner, one of the things I think of is "Do you have the time for me?" because you look at doctors right now, and you can tell they do not have the time to even talk to you. And they're onto the next thing. So if we actually have time, patience with somebody, I think that will be so important. Awareness is vital. What I mean by awareness is to be mindful and aware of not our own mind, not our own thoughts all the time thinking, thinking thinking, but open as reflection to our surroundings. So in the Tibetan practice, the awareness would be analogy would be a mountain lake. A mountain lake when it is glassy and flat without any wind is a reflection of its surroundings. It's like a mirror of its surroundings. When there's turbulence, it's no longer a reflection of the surrounding For our own situation, what this is referring to is that when we are present without so wrapped up in our own thoughts, we then become a reflection of the surroundings we are here we see, we are with the patients, we're listening. And then we can see things that maybe others would miss, because we're aware. So I think bedside manner, time, taking the time to act, and it doesn't actually, it doesn't take any more time. Really, it doesn't take any more time. It just takes us in our minds to say, You have my full, undivided attention. I'm not thinking about other things right now, what is going on? I'm here with you, I want you to be healthy, I love you, I care for you, and to be present. So I think that is what I would describe as bedside manner when I meet with somebody who is showing me that they're going to take the time to sit for two seconds and be with me, because you could be with me for five minutes, but not be with me. versus being with me for 30 seconds and actually feel it. So that's what I would say bedside manner is."* (Subject 5: 1:08:51)<br><br>*"Predisposition is sometimes discussed within Western medicine in a manner that actually dismisses the impact of humans. Like racism, black mothers and death/mortality rates, etc. But does not focus on the impact from other humans: impact of being enslaved and that stress impact for generations, etc. Seems like a lot of emphasis on the focus of 'here and now' of predisposition without really discussing/honoring the past wounds that led to the now."* (Subject 3: 17:24)<br><br>*"Western medicine is very colonial – 'scientific' discounts a lot of important science and negates the relevance of other systems, which means we are missing out on some important opportunities that impact medicine and public health.* (Subject 3:7:00) |

**PRENATAL ESSENCE**

**Definition:**

Accumulation of contextual and physical influence from parents, fetal experiences while in utero, and generational factors; fixed at birth.

**Related Codes/Themes:**

- Genetics
- Access (personal + generational)
- Generational Trauma

*"Actually, in the older texts, they would say that the prenatal you really can't, it's generational. You can't …change. And it's, but now you're, you know, maybe we can change it with genetics, the genetic manipulation, we can do that."* (Subject 12: 6:29)

*"Jing that that gene is never isolated from the three treasures"* (Subject 12: 51:49)

*"Humans are made to adapt. Right? So there's always adaptations to our environment, right? What we don't recognize is whether that adaptation happens within the natural environment or within manmade structures, right? where people are either allowed to live free or live within oppressive societies or structures, right? The changes or the adaptations that happen, really come through. This… is a challenging subject, because this is a perfect example where mess Western medicine just continues to ignore chronic issues, right. One of the one of the most telling issues is when we look at the impact of stress on you know, mammals, right? They did. They did a study on the infant mortality, infant mortality rates, and pregnancy mortality rates of women, black women in particular*. (Subject 3: 12:46)

*"However, I will say that based on what I've learned about trauma, and the impacts of trauma, *(slight pause)* trauma is real. And the impact of trauma is real. And it can be, it can influence generations. So the trauma that-that one experiences could impact one's offspring, even if that offspring hasn't experienced the same trauma. So we talk a lot about generational trauma. And there's a lot of talk about, like, really, probably- really holistic care for children doesn't start when they're born, it starts when the mother is pregnant, because that whole experience in the womb has an impact when the child is born."* (Subject 4: 12:02)

*"So pre Heaven is kind of… is the kind of coming of our ancestors together to create. So it's not just it's not just the mother and father, but it's the whole ancestry all coming together, in my mind. "* (Subject 6: 6:09)

*"Pre-heaven essence captures the idea of energetic potential – this concept does not necessarily get captured that way within Western medicine."* (Subject 7: 12:25)

*"Pre-heaven essence/jing is very hard to deplete; can deplete post-heaven jing. Very hard to distinguish the two, so I don't. "*(Subject 1: 11:47)

*"So it's responsible for growth and maturation and reproduction and a whole bunch of things. Right? It's the strength of the constitution. It's..our Western genetic heritage, if you will."* (Subject 6: 5:15)

*"At birth, for a vaginal birth, the child is colonized with the mother's vaginal flora which colonizes their gut for digestion. Right. So if we think about it, in the terms that we're talking now, that's the mother's mother's mother's mother's mother's mother's mother's, Vaginal flora. So, um, like I have this case right now with this child has severe eczema. And it was in Europe, for C sections or C births. If the vaginal flora is healthy, if there's no strep B, or HPV or any other disorder, they*

| | |
|---|---|
| | *actually do a swab inside the cheek, the neck, under the arms, and the groin, the inguinal ligament area. And they found that there are less food allergies when they do that. Now we don't do that in America, because I don't know why they do it, but they don't. So this child, what's interesting is the mother is Filipino, and the father is Colombian. So it's this beautiful kind of cultural mix. So we had the baby, the mom is cooking the baby traditional Filipino food, and we did do a vaginal swab. Right, so the eczema started clearing because that Flora changed, right? And so that kind of process of honoring that kind of millennial, a millennia millennia of birth, and carrying on, has allowed this five month old baby to kind of reset. Yeah. You're talking about put the influence of post heaven.”* (Subject 6: 16:55)<br>*“Yeah, it's what we do know that the carrier mom, the matriarchal carrier, I think is the term influences the microRNA of the fetus, which changes their post heaven.”* (Subject 6: reference example at 46:59) |
| **POST-NATAL ESSENCE**<br><br>**Definition:**<br><br>Accumulation of contextual and physical influence from parents/caregivers, patient experiences, and generational factors post-birth.<br><br>**Related Codes/Themes:**<br>● Epigenetics<br>● Fertility<br>● Creativity<br>● Metabolism/Digestion*<br>● Yangsheng*<br><br>**Discussion:**<br>A particular phase of time after birth.<br>The beginning portion of this phase is completely dependent on caregivers to provide access to postnatal qi development, education, and access which lay the foundation for adolescent, teen, and adult developmental life for post natal existence.<br>Throughout the lifespan of an individual, a practice of self cultivation. | *“ (fertility): “So when I'm reading the face, and I see someone's fertility, I'm looking at a really small area. I'm looking right here, I'm looking to see how wide the philtrum is, how long the philtrum is, how deep the filter is, and looking to see, you know, whether or not there's the ability for the body to, to get pregnant, to hold that pregnancy to maintain that pregnancy. It's something that's really, really important. And I think that …what’s the word that I want to use…, there's a lot of..... a lot of signs that say that fertility is compromised by the ways that we live. The ways that we live right now. And so there's some, some really interesting ways of handling things about Jing that say, Look, I'm gonna take care of myself, I'm going to stop working, or I'm going to stop, you know, doing as much, I'm going to work part time, and whatever it takes, you know, but some people, of course, have work and be fully functional. So it really depends on what people need. And so the idea is, how important is, you know, fertility? And are you willing to compromise a little bit in order to have a baby, you know, so? “* (Subject 2: 12:28) |

| | |
|---|---|
| **DETERMINANTS OF HEALTH**<br><br>**Definition:**<br><br>The factors that interplay and create multi-dimensional impact on the mental, emotional, physical and spiritual health of an individual.<br><br>**Related Codes/Themes:**<br>● Barriers<br>● Multi-Faceted<br>● Epigenetics<br>● Religion<br>● Zip Code<br>● Environmental Stress<br>● Nutrition<br>● Sleep<br>● Fear/Stress<br>● Financial Health<br>● Socioeconomic Status<br>● Race<br>● Education<br>● Trauma<br>● Objective Indicators of Health<br>● Data* | *"Yeah, I, I think that if you were born in a difficult situation with regards to the location that you're born into, and with your abilities from that location, if you're born in a very tough inner city neighborhood, for instance, then that is so hard to break out of from a generation standpoint, because of education. The education situations, there's such a disparity, especially now in the pandemic, from one area to another, my wife works at an inner city school in Providence. And there was a student who couldn't get online because she couldn't get her computer to connect, and she didn't know what to do. So during the pandemic, my wife actually had to go visit her house, now it's a large family in a tiny apartment. And they open up the laptop, and the girl says, See, I don't know, I type in the address and it doesn't work. And then my wife realized, you don't have an internet, they didn't have any Wi Fi, they had to get a girl a hotspot and set her up the school had to do this. So what a disadvantage here is someone that has no internet, and they're living in a situation mom and dad are away, mom and dad have to go work just to make ends meet. So there's not a way for her to be able to. And this impacts nutrition, too. So environmental nutrition and finances, I mean, those are really important things for our overall well being in our health. And so in that situation, nutrition is not as healthy. So the schools were delivering the box lunches to the kids that were on that program, then providing the internet, but then you look at another situation, on the other end of the spectrum. And kids have everything, they've got all the things that they need at their disposal to be able to be successful. But that doesn't guarantee success because like I was saying before, from the karmic situation, it could add to complacency. One of the things I've seen with the college students is that mom and dad have done so much, they've done too much. So now you don't know how to operate on your own. And you don't know how to navigate the world with other people who are from different cultures, because you have been isolated in a bubble. And that can also add to issues. So I see that getting out. So from the highest level, generation and generation that wealth is getting passed down. For the most part. I mean, some are choosing not to do that, but many are and if you're not passing the wealth down, for instance, then that will the child will still have the higher level of education likely and the opportunities to be able to get into the schools and do those things so that they can be better off. So I think that leads to also help because there's a direct correlation to where we live and our overall well being and health and there's a correlation to our economic status as well. And then from the other side of it and family. Are you hard if you don't have all of the needs, like having the internet very hard to be able to get out? And I'm looking at sending my oldest to college next year? Well, it's so damn expensive. Like, how does anybody afford that? Right? So there is this perpetual wheel of motion that's in play and very hard that's going from generation to generation. And it does impact the overall health status from one generation to another. Also, the family is the parents, whatever their behavior may be, is going to impact the child's. If the parents are not healthy, and don't have healthy activities. And lifestyles, then likelihood is a child's goal. Follow along those lines hard to break free from that, too. Not impossible, but difficult."* (Subject 5: 51:43)<br><br>*"The impact of trauma is real. And it can be, it can influence generations. So the trauma that that one experiences could impact one's offspring, even if that offspring hasn't experienced the same trauma."* (Subject 4: 11:18).<br><br>*"Lifestyle enhancers are very appropriate. I, I've always thought that* ***Yang Sheng*** *is very, very necessary for creativity, as well as fertility" "Yes, and there are things* |

| | |
|---|---|
| | *that the way that you eat, there's things that the way you sleep, there's things about all different aspects of life, and I think they're really critical. And I also think it's really important to recognize that, that we're looking at other scholars like you, um, Sung Sing Yao, is big the writer on Yang Sheng.”* (Subject 2: 8:00 +10:25)<br><br>*“However, what you’re describing are principles used in Western medicine, like looking at how, for example, trauma like pre-birth trauma, either while you’re in utero or even the trauma that previous generations have experienced, continue to impact future generations, like there’s documented research on that.”* (Subject 4: 7:46).<br><br>*“Ace Scores. Poverty and trauma are the top two negative factors.”* (Subject 7: 37:22)<br><br>*“Stress of money and time impact both the upper echelons of society and low income.”* (Subject 1: 20:54)<br><br>*“Same factors listed as positive or negative – just switched due to access”* (Subject 4: 30:55)<br><br>*“Patient’s self-report of health is on the list of how to identify the state of health. Some things aren’t going to show up on an MRI (or access to MRI) but they are feeling something. Not the best use of science of knowing if you have a gene because it doesn’t actually determine the health outcome, so many other things determine health outcomes (again, genetic roulette of what is expressed).”* (Subject 7:24:36)<br><br>*“We know that zip code is one of the best predictors above all else of your health outcomes and your educational attainment. Zip code of birth is still one of the strongest predictors of long term health outcomes.”* (Subject 7:19:41)<br><br>*“So as you said, for yin and yang it divides into two parts and as you are discussing for jing that is also coming from various parental jing. But also, there is like post parental jing, which is going to be impacted by your- what you eat, nutrition, or health care status.”* (Subject 11: 6:22)<br><br>*“Truly unconditional love can be a protective/positive factor.”* (Subject 7: 42:57)<br><br>*Economics, right, we're really talking about we're really talking about the, the 'haves' and the 'have nots', right, the actual distribution of wealth, right, and where it tends to center, and where people end up in the margins, we forget that there are structures as early as the 17, and 1800s, that still have impact that still sits in the brains today, right, except we call it implicit bias. And we don't recognize its roots.*<br>(Subject 3: 30:51)<br><br>*“ We know that zip code is one of the best predictors above all else of your health outcomes and your educational attainment. Zip code of birth is still one of the strongest predictors of long term health outcomes.* (Subject 7: 19:41) |

| | |
|---|---|
| | *"Patient's self-report of health is on the list of how to identify the state of health. Some things aren't going to show up on an MRI (or access to MRI) but they are feeling something. Not the best use of science of knowing if you have a gene because it doesn't actually determine the health outcome, so many other things determine health outcomes (again, genetic roulette of what is expressed." (*Subject 7: 24:36)<br><br>*"Tests for health: sleep quality, range of motion, quality of tissue, etc. Quantity of sleep too."* (Subject 7: 31:40)<br><br>*"Main stressors of his current patients: money and time. Not having enough of either one or the other. Upper echelon of society = not enough time. Lower echelon of society = not enough money. Overall, the stress for either part of society is real and has an impact on their health/well-being."* (Subject 1: 20:54)<br><br>*"Yin/yang within bedside manners. A healthcare practitioner who is warm, friendly feels yang. A healthcare practitioner who is blunter with their boundaries and authority feels yin.* (Subject 11: 48:28)<br><br>*"Western medicine is very colonial – 'scientific' discounts a lot of important science and negates the relevance of other systems, which means we are missing out on some important opportunities that impact medicine and public health."* (Subject 3: 7:00)<br><br>*"Predisposition is sometimes discussed within Western medicine in a manner that actually dismisses the impact of humans. Like racism, black mothers and death/mortality rates, etc. But does not focus on the impact from other humans: impact of being enslaved and that stress impact for generations, etc. Seems like a lot of emphasis on the focus of 'here and now' of predisposition without really discussing/honoring the past wounds that led to the now.* (Subject 3: 17:24)<br><br>*"We're not just scientific people – we are beings of faith and belief as well. But right now, it's still very Christian-centric concepts so things like energy, which is found within African culture, is being overlooked."* (Subject 3: 9:26) |

#### Table 3: Acronym Key

Below is a table of acronyms used throughout the text

| ACRONYM | EXPANSION |
|---|---|
| BM | Biomedicine |
| CDC | Center for Disease Control |
| DOH | Determinants of health |
| EAM | East Asian Medicine |
| SDOH | Social Determinants of Health |
| WHO | World Health Organization |
